# Technology Enabled Community Outreach to Achieve Large Scale Coverage of Family Planning Services in Urban Pakistan: Implementation Results from the Aapi Model

**DOI:** 10.64898/2026.03.07.26347840

**Authors:** Syed Saddam Haider, Heena Tariq, Muhammad Ibrahim, Wahid Husain, Aisha Tauqeer, Muhammad Imran Javed, Ayesha Khan, Adnan Ahmad Khan

## Abstract

**Background:** Pakistan’s fertility rate remains among the highest in South Asia, while its progress toward FP2030 goals has been slow. Poor urban populations, which comprise nearly a third of the country’s population, are often missed by conventional health and family planning service delivery systems. The Akhter Hameed Khan Foundation, with funding from the Punjab government, had demonstrated a community outreach model where family planning access was increased for 278,000 population. The current paper describes its scale up to expand coverage to an additional 800,000 population, through community outreach by local women to adapt to the local context and technology to ensure near 100% coverage, enhance the quality of monitoring and lower the costs.

**Intervention:** This digitally monitored, community-led outreach model (“Aapi”) was scaled across 23 urban and peri-urban union councils of Rawalpindi District between November 2022 and December 2024. Locally recruited female outreach workers (Aapis) conducted household mapping and counseling, provided short-term FP methods, and referred clients for long-acting methods. Real-time dashboards, GPS verification, and automated data checks enabled continuous supervision and adaptive management.

**Implementation Results:** The program registered nearly 100,000 married women of reproductive age, achieving near-universal coverage in the intervention area. Contraceptive prevalence rose from 36% to 45% within two years, and 37% of short-term users adopted long-acting methods. Average implementation cost was PKR 1,981 (US$7.10) per user - less than half that of comparable national FP outreach programs. Digital monitoring helped improve data completeness, worker accountability, and program efficiency.

**Lessons Learned:** Embedding digitally supported outreach workers that are from the community in urban neighborhoods can achieve universal FP coverage at low cost. Key enabling factors included local recruitment, simplified digital tools (including digital automation), frequent feedback loops, and flexible supervision. Challenges included staff attrition and sustainability of incentive mechanisms. The Aapi model describes a feasible, scalable approach for improving FP access and accountability among underserved urban populations in Pakistan and similar low- and middle-income country (LMIC) settings.

## INTRODUCTION

Pakistan’s population of 245 million continues to grow at annual rate of 2.54%,^1^ despite decades of investments in family planning (FP) infrastructure. The country is struggling to meet its FP2030 target of 50% contraceptive prevalence rate (CPR) by 2025 (or 60% by 2030),^2^ and its fertility rate of 3.55 per women outpaces its neighbors, India (2.1), Bangladesh (2.05) and Iran (2.15).^3,4^ High fertility and a growing population have contributed to Pakistan’s low Human Development Index ranking of 161, well behind its regional peers.^5^

Several reasons account for this poor performance. In the traditional clinic-based approach, the catchment of one or few isolated clinics serves too few women in the locality to be measured in national surveys.^6,7^ In Pakistan, only the government’s Lady Health Workers (LHWs) program reached population level impact in Pakistan.^8,9^ Private sector programs such as the Sukh Initiative, Marie Stopes Society, the Health and Nutrition Development Society (HANDS), and the Akhter Hameed Khan Foundation (AHKF) have shown promise, albeit at limited scale.^10,11^

Another key gap is that although urban poor account for 30% of the population, they receive few public services due to undercounting and political neglect.^7,11^ Globally, community and outreach-based interventions have brought local knowledge into addressing both demand and supply side components of FP.^12–14^ In Pakistan, lady health workers did so rural populations, but largely did not reach the urban poor.

Other factors such as insufficient supplies, inconsistent programs and limited demand from women that aren’t empowered and men that aren’t engaged in the FP dialogue, along with missed opportunities of postpartum family planning further limit effectiveness of FP efforts.^2^ Finally, there are few examples of using technology in Pakistan to improve coverage, ease programming, reduce costs or improve efficiency.

Previously, AHKF, with funding from the Punjab provincial government, had demonstrated a successful test of a women-driven hybrid (demand + supply), community-based model to rapidly increase in CPR by 11% among 278,000 population (36,000 married couples) at low cost in Dhok Hassu, an urban informal settlement in Rawalpindi, Pakistan.^11^ The current paper explores the effectiveness and feasibility of adding technology to scale that model to an additional 0.8 million population (100,000 married couples) across 23 Union Councils (UCs) of Rawalpindi District. It describes service delivery, community engagement and technology use that allowed the project to cover such a large population while demonstrating near 100% coverage of the intervention so that elements of the intervention may be replicated.

## METHODS AND IMPLEMENTATION APPROACH

This paper presents a descriptive implementation study of the Aapi Model (Figure 1) using routine program data collected between November 2022 and December 2024 (26 months) in western Rawalpindi in 3 tehsils or 23 union councils (Table 1). The study specifically assessed:

1. The design and operationalization of the community-based Aapi outreach model for family planning at near city-wide scale
2. The use of digitization and geospatial mapping to enhance quality and coverage and reduce costs.
3. Lessons and cost estimates for the scale up.

**Figure 1.** Aapi Model.

**Table 1:** Tehsils and Union Councils for the Intervention.

| Tehsil | Union Councils |
| --- | --- |
| Rawalpindi West | Chak Jalal Din, Jirga, Dhok Hassu, Khayaban-e-Sir Syed |
| Taxila | Lab Thathoo, Saray Kala, Jalala |
| Gujjar Khan | City-1, Mandra, Kalyam Awan, Changa Maira |
|  | Badhana, Gulyana |
|  | Rammay, Thathi, Narali, Sahang |

The intervention locality (Table 1) were identified by the funder based on poverty (as defined by the Benazir Income Support Program [BISP] national cash transfer program, which uses the National Socio-Economic Registry [NSER] and specifies a score of 32 [scale: 0–100] as the cutoff for absolute poverty; https://www.bisp.gov.pk). Community was mobilized and local women called Aapis (sisters) conducted systematic outreach to married women of reproductive age (MWRA, 15–49-year-old married women) through household visits. Aapis were selected from local communities, and to address limitations on women’s mobility in these communities, they work within a few streets of their homes. Funding was provided by the Punjab Population Innovation Fund (PPIF) of the Punjab government and was implemented by AHKF.

### Reach and Target Population

To achieve nearly 100% coverage and avoid exclusion of any household in the intervention area, particularly due to poverty and marginalization, each locality’s population was estimated using Google Buildings^®^ data. Google Buildings is open-source data that identifies the outlines and heights of each building in the area which is used to estimate local area population. The process was validated within ±9% of census counts and allows all houses to be marked with Google Plus Codes®, making it possible to revisit homes in urban slums where houses are seldom numbered. Out of the 115,000 mapped households 99,956 were covered by the intervention. In 2022, we were unable to discern residential buildings from commercial buildings in Google Buildings data, which accounted for the additional households in the estimate. By December 2024, 99,956 households visited represented around 98.5 of all households in the intervention area.

### Community Mobilization and Workforce Recruitment

Forty outreach workers (called Aapis or elder sisters) and six men were selected from local communities, on expressed self-interest (motivation), minimum grade 8 education, availability of 4-5 hours of time, willingness and ability to visit community households, and permission from the head of their household to engage with AHKF.

Aapis are trusted community members, chosen for their familiarity with the local population, that allows access to the community and understanding of local context and also served as central nodes in a local network that also includes local influential people, community groups and local healthcare providers to fully ground the intervention in the community.

Aapis initially received a monthly stipend of PKR 8,000 (USD 28) that was increased to PKR 10,000 (USD 36) in July 2023 to address inflation. Typically, Aapis worked flexibly for 2-6 hours a day, visiting up to 10-12 households daily, for 5-6 days per week. Aapis supplemented their health visits with active BiB sales, part-time work as polio or other government campaign workers, and through paid referrals to local private providers.

Aapis carried a **Business in a Box** (BiB) with short term methods (condoms, pills provided by the Punjab government population welfare), and some common use household products and women-centric supplies. They received a 12-month revolving credit of PKR 20,000 (USD 71) to purchase products and had the autonomy to determine pricing and contents they carried.

### Implementation Process

Selected Aapis received 3-week low-literacy, pictorial based, interactive training modules on health counseling for maternal-reproductive health, immunization schedule, side effects, nutritional growth monitoring, enterprise building, communication and marketing. Each group of Aapis had a cluster lead (supervisor) that served as a technical resource and was from the same community. They were also assisted by 5 male activists, providing support to Aapis as required.

Aapis registered each new household, they collected basic, FP and RH information on a form designed as a “nudge” to initiate a conversation about FP needs. It also limited Aapis – who share common community biases from – from opting out of having these conversations or ignoring certain households, such as newly married couples and those with few children from these conversations. For existing users, Aapis identified the current method and inquired if they wanted to continue or switch to another method. In either case, the conversation then led to available options for FP, such as condoms and oral pills, which were given right away, and referrals for injections, IUCD, implants or contraceptive surgery. Paper-based IEC material was provided to support the counseling, including for non-users. Those who were interested were given contraceptives or referred as above, while those initially uninterested are revisited after three months with tailored counselling sessions.

### Service Delivery and Referral System

After the initial interview, Aapis provided doorstep FP counseling, short-term contraceptive methods (from the Punjab Population Welfare Department) and referrals for long-acting reversible contraception (LARC) (to local government and private providers). They also identified and referred those with severe malnutrition and missed doses of immunization to the local government clinic. In some a proportion of households Aapis also conducted follow-up visits within one week to 3 months after the first registration visit to ask about the experience and any side effects.

### Data Collection and Monitoring Progress

Monitoring was integrated into implementation through a combination of digital tools, in-person validation, direct supervision and structured feedback. At the household visit, Aapis intake interviews were recorded digitally using the SurveyCTO^®^ software installed on the tablets and smartphones provided. The data intake form was designed as a “nudge” to help MWRA review their reproductive preferences and to think about FP choices, since previous experience had shown that counseling provided by Aapis is limited due to their own biases and comprehension.^11^

Digitization helped ensure accurate data entry, for example, by mandating a complete household roster and all essential questions by restricting skipping of relevant answers. The database fed into a Google Sheets based dashboard where quality checks were automated and visualized through a monitoring dashboard that flagged inconsistencies and unusual patterns in the data. For example, pre-designed quality flags identified issues such as data collected at wrong locations, short or inordinately long visits, unusual (e.g., 20 children) or inconsistent answers, too many skips (for example, if too many MWRA for an Aapi report no children) etc. Additionally, a 180 second audio recording would randomly turn on to monitor whether the Aapis were actually asking the questions while filling the form. Referral slips from health facilities were collected and entered manually into the same database. All unusual answers plus a few random selection of acceptable entries were validated by in person visits. The dashboard also embedded location heatmaps to ensure coverage by neighborhoods. It showed granular results for each Aapi, by each locality, and for each day (Figure 2) and depicted visits, current contraceptive use and progress for each individual MWRA served. Results were shared with the program team and cluster leads each week. Cluster leads met weekly with Aapis to review field performance, address challenges, reinforce key performance indicators to address quality issues, visiting missing households or neighborhoods and to review if certain MWRA needed counseling to change a method, for example, someone who had previously accepted a short-term method was offered the choice of a LARC or injections. The use of digital data that was tracked on a geospatial map reduced the amount of field monitoring required, allowing only 2-3 monitoring officers to validate data as above.

**Figure 2.** Data Monitoring Dashboard.

**Figure 3:** Current user complete output flow.

### Analysis Plan

The data were cleaned and prepared for this study in STATA software version 16.0 (StataCorp, College Station, Texas). Duplicate entries of the respondents were excluded from the final dataset based on a unique code generated from respondent name, union council name, date of birth and child number in the family. Descriptive statistics were calculated for continuous variables. Where the data was normally distributed, frequencies with percentages were calculated for all categorical variables and a regression analysis – logistic analysis to analyze the effect of different demographic variables on program outcomes. Furthermore, this paper also evaluates sustainability by assessing the financial and administrative feasibility of program execution for further scaling up.

The analysis framework was based exclusively on programmatic data collected through routine service delivery processes. No separate baseline or endline surveys were conducted by the implementation team due to budget constraints, as the intervention design emphasized continuous engagement and real-time data collection through digital tools rather than isolated evaluation points. The donor commissioned baseline and end line surveys but without control locations and did not share the sampling methodology or results until well after the completion of the project. Instead, the first visits by Aapis were taken as baseline, capturing initial information on demographics, reproductive health status, FP knowledge, access, barriers, and intentions. Similarly, the doorstep service availed at the last outreach or referral visits was considered as endline data.

A detailed registration questionnaire was developed covering MWRA socioeconomic and demographic characteristics, contraceptive usage patterns (current, ever, and never users), contraceptive method preferences (LARC and Short-Term Methods), and willingness to convert from short-term to long-term contraceptive methods. Questions were translated into Urdu to ensure Aapis’ comprehension and maintain high-quality program data. Similarly for the follow and outreach data was gathered using more structured questions similar to registration ones. A detailed Analysis plan has been crafted to which primarily focused geographic coverage, demographic profiling, baseline CPR, after intervention CPR, method mix (current), follow-up coverage, cost effectiveness.

### Cost Calculations

To calculate the cost-effectiveness of the intervention, costs per household reached and costs per FP user were assessed as key metrics. We used overall program costs that included salaries, training, technology, transportation and other management fees. Costs of commodities were not included as these were provided for free by the government.

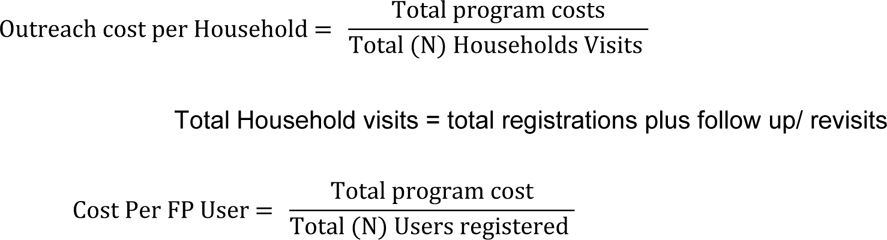

### Qualitative Analysis

Qualitative analysis assessed uptake of counseling, performance of the Business in Box (BiB) initiative, work issues of Aapis, provider engagement, to learn how the intervention was operationalized. For most part this included complementary operational progress quality assessment with managers and Aapis about their work and excerpted from notes of weekly and other meetings.

### Role of Government in Providing Supplies

The Punjab Population Welfare Department (PWD) provided contraceptive supplies such as pills, condoms, and referrals for IUDs at their local clinics. The Punjab Population Innovation Fund (PPIF, also the funder) provided program oversight.

### Ethical Oversight

The study was conducted under the oversight of the Punjab Population Innovation Fund, a public service company of the Punjab government. The study protocol, including the verbal informed consent by MWRA at each interview, was approved by the IRB of Research and Development Solutions, which is registered with the US NIH Office of Human Research Protection (IRB00010843).

## RESULTS

### Reach and Population Profile

A total of **99,956** MWRA were reached during this (second) phase of the intervention from across 3 tehsils, Rawalpindi (42,981, 43%), Gujjar Khan (28,987, 29%), and Taxila (27,988, 28%). The mean age of the MWRA was 32 (SD ± 7) years, with 21% in the 25 to 29 years old age group and another 23% between 30 to 34. Only 9% reported working for an income. On the contrary, 97% of spouses were employed (Table 2).

**Table 2:** Demographic characteristics of MWRAs in three tehsils of Rawalpindi district.

| Characteristics | MWRA<br>n (%) |
| --- | --- |
| Total MWRA reached | **99,956** |
| Tehsil |  |
| Rawalpindi | 42,636 (43) |
| Gujjar Khan | 28,866 (29) |
| Taxila | 28,454 (28) |
| Age groups distribution |  |
| 15 to 19 | 1,694 (2) |
| 20 to 24 | 13,666 (14) |
| 25 to 29 | 21,412 (21) |
| 30 to 34 | 22,729 (23) |
| 35 to 39 | 21,240 (21) |
| 40 to 44 | 11,842 (12) |
| 45 to 49 | 7,373 (7) |
| Women's Employment | 8,756 (9) |
| Spouse's Employments | 96,918 (97) |
| Children (any) | 80,792 (81) |
| Less than 3 children | 29,564 (37) |

### Baseline Contraceptive Use at Initial Registration

At registration (baseline), 35,777 (36%) MWRA reported using some form of family planning (CPR: 36%, modern CPR [mCPR]: 31%) with the highest usage in Rawalpindi (9,542, 54%), followed by Taxila (4,460, 26%) and Gujjar Khan (3,481, 20%). Most existing users (73%) were 30 years or older, consistent with national data that FP starts well after fertility peaks between ages 25-29 years. Younger women under 25 constituted only 8% of existing users but 21% of never users. Among lapsed users, women over 40 represent nearly 30%, suggesting discontinuation later in reproductive life.

Condoms were the most common method at registration among both current (43%) and lapsed (those that had used FP previously but had since discontinued) users (39%), followed by oral pills and tubal ligation. Among lapsed users, injections (19%) and IUCDs (9%) accounted for a significant minority. Traditional methods accounted for approximately 12% in both groups.

**Table 3.** Age Distribution of Existing Users.

| Age Group | All MWRA | Current Users<br>N (%) | Lapsed Users<br>N (%) | Never Users<br>N (%) |
| --- | --- | --- | --- | --- |
| 15–19 | 1,694 (2%) | 203 (0.6%) | 36 (0.5%) | 1,455 (2.6%) |
| 20–24 | 13,666 (14%) | 2,548 (7.1%) | 481 (6.3%) | 10,637 (18.8%) |
| 25–29 | 21,412 (21%) | 7,082 (19.8%) | 1,418 (18.7%) | 12,912 (22.8%) |
| 30–34 | 22,729 (23%) | 9,122 (25.5%) | 1,762 (23.2%) | 11,845 (20.9%) |
| 35–39 | 21,240 (21%) | 9,387 (26.2%) | 1,607 (21.2%) | 10,246 (18.1%) |
| 40–44 | 11,842 (12%) | 5,113 (14.3%) | 1,268 (16.7%) | 5,461 (9.6%) |
| 45–49 | 7,373 (7%) | 2,322 (6.5%) | 1,017 (13.4%) | 4,034 (7.1%) |
| **Total** | 99,956 (100%) | 35,777 (100%) | 7,589 (100%) | 56,590 (100%) |

**Table 4.** Method Mix of Existing and Lapsed Users at Registration.

| Method | All Ever Users<br>N (%) | Current Users<br>N (%) | Lapsed Users<br>N (%) |
| --- | --- | --- | --- |
| Condom | 18,283 (42%) | 15,298 (43%) | 2,985 (39%) |
| Pills | 5,247 (12%) | 4,365 (12%) | 882 (12%) |
| Injection | 4,466 (10%) | 3,020 (8%) | 1,446 (19%) |
| IUCD | 2,528 (6%) | 1,877 (5%) | 651 (9%) |
| Implant | 589 (1%) | 453 (1.3%) | 136 (1.8%) |
| Tubal Ligation | 5,933 (14%) | 5,525 (15%) | 408 (5.4%) |
| LAM | 883 (2%) | 776 (2.2%) | 107 (1.4%) |
| Traditional | 5,347 (12%) | 4,394 (12%) | 953 (13%) |
| Other | 90 (0) | 69 (0.2%) | 21 (0.3%) |
| Total | 43,366 (100%) | 35,777 (100%) | 7,589 (100%) |

### Program Supported Uptake

Once they were visited by an Aapi, 34% of MWRA chose to receive condoms or pills (15,772 – 16%) or a referral (18,623 – 19%) from an Aapi. Of these, 65,561 (66%) chose not to receive services from Aapis. This included 47% of existing users, while 53% of existing users shifted to services from an Aapi. Importantly, 23% of previous non-users initiated a method with Aapis, 10% of these taking a condom or a pill and 13% a referral. Most of those choosing to work with Aapis were between ages 25-39 years old, while those that opted out were evenly distributed across all ages.

Among those that chose not to avail themselves of services by Aapis, most had availed tubal ligation (33%), followed by traditional methods (20%) and condoms (19%). Among those that chose to work with Aapis, most took a condom (64%) either directly or through referrals while 19% chose pills. Most importantly, 10% of women that worked with an Aapi asked for referrals for injections.

**Table 5:** Program Uptake.

|  | No Interaction with the program | Program Uptake | Took condoms or pills at Doorstep | Took a Referral from Aapis |
| --- | --- | --- | --- | --- |
| **Total** | 16,765 (100%) | 35,777 (100%) | 9,418 (100%) | 9,594 (100%) |
| Current User | 16,765 | 35,777 | 9,418 | 9,594 |
| Ever User | 5,173 | 7,589 | 885 | 1,531 |
| Never User | 43,623 | 56,590 | 5,469 | 7,498 |
| **By Age Groups** |  |  |  |  |
| 15 to 19 | 1,484 (2%) | 1,694 (2%) | 112 (1%) | 98 (1%) |
| 20 to 24 | 10,780 (16%) | 13,666 (14%) | 1,520 (10%) | 1,366 (7%) |
| 25 to 29 | 13,078 (20%) | 21,412 (21%) | 4,359 (28%) | 3,975 (21%) |
| 30 to 34 | 12,478 (19%) | 22,729 (23%) | 4,509 (29%) | 5,742 (31%) |
| 35 to 39 | 12,423 (19%) | 21,240 (21%) | 3,600 (23%) | 5,217 (28%) |
| 40 to 44 | 8,751 (13%) | 11,842 (12%) | 1,237 (8%) | 1,854 (10%) |
| 45 to 49 | 6,567 (10%) | 7,373 (7%) | 435 (3%) | 371 (2%) |
| **By Methods** |  |  |  |  |
| Condom | 3,211 (19%) | 15,298 (43%) | 6,975 (74%) | 5,112 (53%) |
| Pill | 719 (4%) | 4,365 (12%) | 1,937 (21%) | 1,709 (18%) |
| Injection | 1,198 (7%) | 3,020 (8%) | 97 (1%) | 1,725 (18%) |
| IUCD | 1,796 (11%) | 1,877 (5%) | 20 (0%) | 61 (1%) |
| Tubal Ligation | 5,499 (33%) | 5,525 (15%) | 4 (0%) | 22 (0%) |
| Implant | 428 (3%) | 453 (1%) | 6 (0%) | 19 (0%) |
| LAM | 476 (3%) | 776 (2%) | 114 (1%) | 186 (2%) |
| Traditional | 3,384 (20%) | 4,394 (12%) | 261 (3%) | 749 (8%) |
| Other | 54 (0%) | 69 (0%) | 4 (0%) | 11 (0%) |

### Method Switching/ Program Pathway

Method mix changed considerably with the intervention. Nearly half of all existing condom and pill users opted to work with Aapis, essentially receiving free supplies. Of most significance are the 5,723 MWRA that received referrals for injections. Around half of these were previous condom users. Most IUCD users chose not to work with an Aapi. Among the 56,590 MWRA that had never used any contraception, 12,967 opted to initiate a method with Aapis, 58% of whom received a referral and 32% supplies at the doorstep. Injections were the most referred method (3,799, 29%), followed by IUCD (2,880, 22%). Condoms were the most common method for doorstep delivery (3,967, 31%).

### Predictors of FP Service Uptake

We estimated a multivariable logistic regression model to assess factors associated with service status (binary outcome). The model included women’s age, fertility composition (number of living sons and daughters), client type, urban wealth quintile, women’s employment status, husband’s employment status, and child immunization status. All categorical variables were entered as indicator variables with clearly defined reference categories. We found that being in age group 25-34 years was associated with use of services (AOR 1.34-1.39) as was increasing number of children, richest wealth quintile, if the husband worked. Employment of the woman herself was not statistically significant.

### Program Cost

We calculated the cost of the program for each household reached and for each user served. The program grant from the government was PKR 42 million (USD 151,525). In addition, RADS made available personnel for monitoring and research through its corporate social responsibility worth PKR 10 million (USD 36,077).

The calculated cost of outreach per household for the program was PKR 413 (USD 1.49), while cost of the program per FP user for the program was PKR 1,981 (7.13 USD).

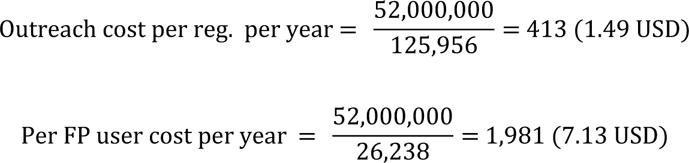

### Barriers to Contraceptive Use

We inquired about the possible barriers to family planning by looking at both reasons for discontinuation among lapsed users and reasons for never using an FP method among never users. Among never-users of FP, 44% said their family was not complete, followed by the husband not allowing FP (23%), fear of side effects (13%), husband being away from home (7%), lack of fertility (6%), and illness (4%). Key reasons for the discontinuation of FP methods were planning a child (37%), followed by side effects (21%), lack of access to a healthcare provider (9%), husband doesn’t allow its use (14%) and no information about FP (8%). Inability to afford any FP method (2%), and nonavailability of preferred method in the market (1%) accounted for a minority of reasons.

### Use of Insights from Data to Guide Programming: Monitoring with Digital Data

Real time visualization allowed rapid responses to programming issues. Data form each Aapi was captured on a tablet or smartphone. The form was designed as a nudge tool based on previous experience that prompted a discussion about the need for FP and about specific methods.^11^ Data were time and location stamped and together with Google Buildings data which allows mobility, could even track how long an Aapi spent working in the field (time spent in client homes plus time spent walking between them). Weekly reports - households visited, their methods taken up, key gaps such households that needed follow up and neighborhoods that were behind - were generated by Aapi and neighborhoods, to track the field team progress on a daily and weekly basis.

Time, duration and location of visits, completeness of sections, outliers among demographics, FP use etc. checks from datasets, along with limited field visits to validate the digital data allowed the monitoring team to identify data quality issues and improved data transparency, while enabling rapid, data-driven course corrections in real time at both the field and management levels.

Several key patterns were observed. There was high initial engagement with follow-up visits that tapered after the second contact, variations in referral uptake across clusters, and inconsistencies in data entry that flagged the need for additional training. It also highlighted peak days and times for outreach activity, enabling better scheduling and resource allocation.

### Qualitative Assessment of the Model

Internal reviews and formal qualitative interviews of project personnel were conducted throughout the implementation period to inform programming. These interviews were analyzed thematically to understand the feasibility, acceptability, and adaptation of FP during the project and for scale up.

A total of 817 counseling sessions were conducted with 6681 participants. However, monitoring revealed that many sessions were not of sufficient quality. They were often rushed, and Aapis tended to be judgmental, focusing more on delivering information than engaging in an interactive dialogue through active listening and exchange with clients. This quality improved only minimally despite remedial training. Despite these challenges, an additional 20 - 30% of non-users expressed interest in initiating family planning.

Aapis are low literate women, most of whom had serious limitations on their mobility outside their homes and had few job or economic prospects. When they are allowed to go out, it is for a few hours and not for long distances. Hence, limiting their work to within a few lanes of their homes made sense. Working on women’s health made it easier for them receive permissions from their homes and doing this work afforded them stature within their communities, where they came to be recognized as role models by other young women.

Low-literate women that became Aapis initially struggled with their own biases against counsel young couples or those with 1-2 children for family planning. This require retraining and values clarification. They also felt challenged when using household forms and BiB inventory tools. Rather than voicing their difficulties, many chose to simply resign or stop attending altogether. It took several months for both Aapis and the project team to fully grasp the model and the expectations associated with their roles. Over time, with repeated small-group refresher sessions, a performance framework was co-developed. Aapis began receiving weekly feedback on their household visits and BiB inventory management, which helped improve their performance, rapport with the community, and counseling abilities steadily.

There were notable contextual differences across implementation areas. In Gujar Khan and Taxila, which are predominantly rural, only local Aapis – as opposed to those that were recruited from a distance - who did not face commuting challenges stayed with the project. These Aapis could access sufficient numbers of clients and households and were generally satisfied with the stipend and the BIB activities. In contrast, in Rawalpindi, although commuting was not a major concern, the availability of alternative opportunities offering higher income contributed to a higher turnover rate among Aapis. Additionally, maturity, prior experience, and family responsibilities of Aapis played a significant role in their retention. Those with greater responsibility and commitment were more likely to continue working with the project.

### Business in a Box (BiB)

Aapis received support with a *Business in a Box* (BiB) initiative as a sustainability strategy. They received training in entrepreneurship and accounts keeping and a starter grant of PKR 10,000 (USD 36) which they used to buy, carry and sell household items such as garments, dry spices, cosmetics etc. – based on their perception of the market need - to generate additional income. This allowed Aapis to add around PKR 2,532 (approximately USD 9.02) a month to their income and 26 Aapis (60%) continued the business even after the closing of the project as a side income stream, while 40% of Aapis struggled to make substantial sales. Interestingly, the ability to make BiB sales performance and how effectively they did family planning (FP) counseling were not correlated. Some Aapis excelled in sales but were less effective in counseling, while others were strong counselors but weaker in sales.

### Aapis’ Performance and Retention

A total of 65 Aapis and 5 cluster leads were engaged through the course of the project, with at least 43 retained at any given time. Attrition was significant initially when it exceeded 20%, before settling down to below 10%, 5–6 months into implementation. This improvement was attributed to regular capacity building, increased community acceptance and recognition of the Aapis, and the introduction of BiB income generation incentives.

### Male Mobilizers

Two male mobilizers (Rahbars) were engaged from the local community. While the mobilizers were able to engage nearly 3,824 men/husbands, their experience highlighted several design issues. Male mobilizers were even more reluctant than the Aapis to counsel newly married/younger couples. Additionally, men in urban slums work one to two jobs and were not available during daytime working hours. When they were available, they would rather not talk about family planning. Men prefer discussing financial issues, income generation, employment, and local municipal issues. Any discussions of FP had to be embedded in these conversations and supplemented with in person private or phone counseling that some men requested. The inability to fully guide these discussions was related to the skills of male mobilizers. For most part, these men had not worked previously in community mobilization. This was during the pilot study and led to several improvements during the scale-up of the intervention. Mobilizers received enhanced training in group facilitation and positive deviance techniques; session times were extended into evenings and weekends to reach working men; entry-point activities were expanded to include men work break times and municipal issue forums; and follow-up counseling was delivered via phones calls for those who expressed interest.

### Healthcare Providers (HCPs)

A total of 30 (27 private and 3 public) - all women - healthcare providers (HCPs) were trained to provide family planning (FP) and related services. All received training and certification in clinical management (managing referred patients and clients) and infection control and prevention (ensuring the standard procedures that when dealing with referred clients and general patients) for LARC services. Cluster leads provided ongoing quality improvement support, including infection control guidance and IEC materials. Contraceptive supplies were made available at a subsidy (through bulk purchases by AHKF).

Aapis referred MWRA to HCPs for LARCs and injections. This model helped increase HCP client volumes for FP and maternity services, leading higher incomes without requiring direct payments from AHKF. Providers also participated in monthly health camps with AHKF support which further promoted them to the community.

As a result, HCPs saw an average increase of 20–30 new FP clients and over 40 non-FP clients per month - representing a 25% and 40% growth compared to pre-program levels. This contributed to building a sustainable ecosystem for FP uptake through demand generation and strengthened healthcare networks.

## DISCUSSION

This experience adds to the community outreach literature by showing that near-universal coverage can be operationalized and monitored in informal urban settings using low-cost geospatial enumeration and exceptions-based digital supervision to potentially reduce the equity gap that arises when programs reach only the most accessible households – all at a fraction of current costs. The low-cost, community-based intervention, “the Aapi model,” was introduced in 2019 in a low-income urban community of around 280,000 people in Dhok Hassu, Rawalpindi, Pakistan by the AHKF. Following this successful first phase,^11^ the model was scaled up to three additional tehsils of Rawalpindi district in 2022 with a population of 800,000. The current paper describes this scale-up, where contraceptive use increased from 36% at baseline to 45% over two years, including enhancement of LARC uptake from 14% to 22% and 37% of current users transitioned from short-term FP methods to LARC or injections. This model was cost-effective, at USD 7.13 per user and USD 1.49 per household reached annually.

A key facilitator of the success of this model was the mobilization of communities, working with local women to provide household outreach to their neighbors, many of whom are restricted from seeking out health and FP services. The work of low-literate outreach workers was facilitated by using open-sourced technology. We used Google Buildings to identify and count area population and marked each household with a Google Plus Codes to ensure repeat visits in communities where houses are not numbered. This allowed us to reach nearly 100% of the households in the community, with near-zero exclusions. We used digital data capture and backend analytics to monitor progress at lower costs and greater transparency.

### Doorstep Counseling and Referrals with Local Women as Outreach Workers

Global literature shows that culturally proximate female outreach workers improve trust to increase access to and availability of reproductive health services and commodities for women that are otherwise excluded from accessing healthcare services due to limited mobility or gender norms.^10,15–17^ This access is further restricted by low mobility of women in Pakistani communities, particularly in urban slums, where only one in four women feel comfortable leaving their home without permission or chaperone.^11^ The Lady Health Workers (LHWs) program in the early 1990s addressed these gaps to significantly improve FP use, albeit mostly in rural communities, leaving unmet need in poor urban communities.^8,18^ The Aapis model is a proof of concept that the idea works even better in urban slums.

The program also describes that a successful referral system can be set up in the community and that some part of this can be through out-of-pocket payments. Globally, more than half of all FP and general medical services in large LMIC are in the private sector (Ibrahim et al, under review and UNFPA 2023). Understanding how to recruit more private providers that serve FP and perhaps their business models may expand the provision of FP services and - at least based on the Pakistani data - be cheaper than resorting to only public sector alternatives.^19^

### Technology Enablers

The project overcame key obstacles with technology. First, Google Buildings open-source data helped estimate the total population of the catchment area to establish a denominator for measuring coverage of intervention. The same technology allowed marking of households with Google Plus Codes for follow-up. This meant that every household in the community could be identified to eliminate any exclusions. Data were collected on phones, minimizing errors and costs. Cleaned data could be followed on digital dashboards and heatmaps to help guide programming in real time and at household level granularity. Some applications of this included the ability to identify and help transition non-users to short acting and then LARC methods using their data and to identify lanes and neighborhoods that needed attention.

A key learning is that because the technology used was based exclusively off the shelf solutions, it reduces costs and avoids the all-too-common obsolescence that plagues custom developed software. In a global world, it also means that our lessons can be easily replicated by someone in a different part of the world with ease and without needing the capacity for software customization.

### Behavior Change and Demand Transition

In our model, nearly half of previous users chose to receive services through our Aapis. Of these, around 60% opted for either an injection or IUCD, both of which are from the private sector. This suggests that there may be a growing nascent demand for FP in these communities. For long it has been questioned whether poor communities in Pakistan or elsewhere can move beyond passively receiving of contraceptives (or vaccines) to actually demanding and procuring FP for themselves. Our experience suggests that there may be a transition or inflection point, or a critical threshold level of mCPR, beyond which this can happen. This is further supported by recent work showing that average CPR in a neighborhood is by far the biggest predictor of FP use, far eclipsing individual factors such education, employment or wealth.^20^ Although this “inflection point” would likely vary by country, cultures and even localities, it would be useful to explore the contours of such a transition, both for the demand for FP for the couple, and for price points at which they would seek to buy different options available to them.

### Program Costs and Its Drivers

Pakistan government-run national LHW program serves around 700,000 women with FP at (inflation adjusted) USD 36 per MWRA per year,^21^ which is higher than global averages of USD 18- 28.^22^ The lower costs of the Aapi model (USD 7 per MWRA) reflects both the urban location with smaller travel distances, a lack of government overheads, and the considerable use of technology to ensure quality at low costs. The cost of FP per user increased by 40% between the two phases, reflecting inflation (which sometime exceeded 40%) and the increased travel distances in Taxila tehsil, where some of the households were located 30-50 minutes from Aapis homes. These changes inform about potential costs drivers when this program is scaled up to other geographies.

### Potential to Expand the Aapis Model for Primary Healthcare

Successful demonstration of the Aapis model to provide universal outreach to all households in a community suggests that it can be scaled to other services. For example, currently a pilot is testing the potential and costs to screen, counsel and refer all members of a community for preventive services that form the core of primary healthcare is underway. The concept can be extended to address vaccine preventable diseases, or to develop surveillance for outbreaks.

### Sustainability

Integrating micro-enterprise with FP outreach helped improve recruitment and agency and kept the costs low and suggested options for replication into government and other models. Another key element of the model was that it was grounded into community mores. Women have low mobility. Aapis were tasked to visit households within a few streets of their homes, engendering confidence for them and their families. The program allowed Aapis to select their own working hours, usually 4-5 hours day, working around their tasks as wives, mothers and homemakers. Finally, Aapis referred to private providers, and received a fee for the referrals. This added to their income while embedding them further into the community healthcare infrastructure.

## Limitations

Given limitations of funding, the program measurement was limited to using service data. No baseline or endline surveys were conducted, limiting the ability to both measure community change in CPR and to ascribe results to the program. Program data were limited in that anyone who opted to not work with AHKF was not followed up. This means that it is uncertain what proportion of 18,221 such MWRA continued their method. Finally, LARC referral was tracked based on client reports and collection of a portion of referral slips from provider. It is possible that not all slips were returned to the AHKF team. However, such reliance on service or supplies data is common in implementation research.

## Conclusion

Despite constraints, and perhaps due to them, we demonstrate a community embedded and technology enabled, feasible, low cost and robust family planning outreach program at large scale (nearly one million population) that also empowers local women as outreach workers. Lessons from this program, its use of technology and low costs can inform government strategies to reduce the costs of expanding lady health workers to reach urban slums. Embedding such models within provincial systems and integrating broader primary healthcare could transform health service delivery for urban poor in Pakistan and LMIC.

## Data Availability

information will only be available after acceptance

## ANNEX 1: ADDITIONAL ANALYSES

### CURRENT USER OUTCOME BY AGE

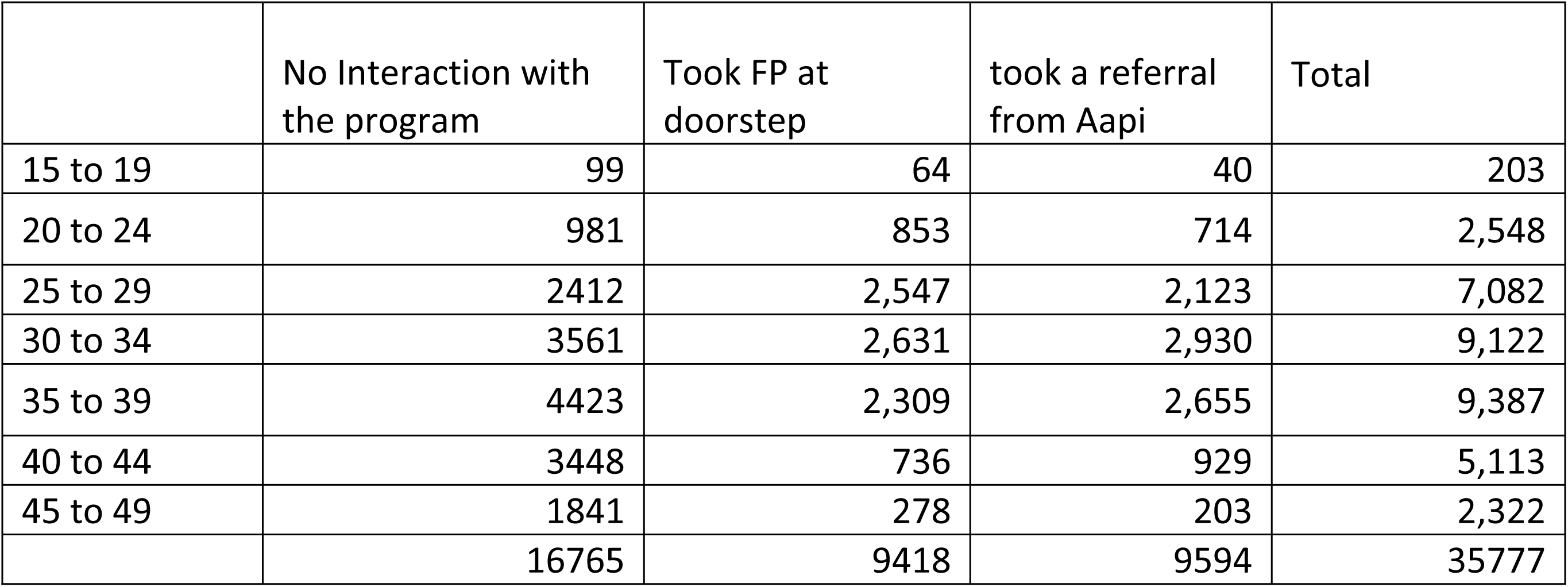

### EVER USER OUTCOME

| Ever User | No Interaction with the program | Took FP at doorstep | took a referral from Aapi | Total |
| --- | --- | --- | --- | --- |
| Condom | 1771 | 493 | 721 | 2,985 |
| Tablet | 486 | 177 | 219 | 882 |
| Injection | 955 | 114 | 377 | 1,446 |
| Tubal ligation | 403 | 3 | 2 | 408 |
| IUCD | 535 | 30 | 86 | 651 |
| Implant | 105 | 10 | 21 | 136 |
| Traditional | 822 | 47 | 84 | 953 |
| LAM | 80 | 8 | 19 | 107 |
| Other | 16 | 3 | 2 | 21 |
| Total | 5173 | 885 | 1,531 | 7,589 |

### EVER USER OUTCOME BY AGE

|  | No Interaction with the program | Took FP at doorstep | took a referral from Aapi | Total |
| --- | --- | --- | --- | --- |
| 15 to 19 | 26 | 5 | 5 | 36 |
| 20 to 24 | 322 | 61 | 98 | 481 |
| 25 to 29 | 931 | 209 | 278 | 1,418 |
| 30 to 34 | 1050 | 245 | 467 | 1,762 |
| 35 to 39 | 954 | 215 | 438 | 1,607 |
| 40 to 44 | 955 | 112 | 201 | 1,268 |
| 45 to 49 | 935 | 38 | 44 | 1,017 |
|  | 5173 | 885 | 1531 | 7589 |

### EVER USER COMPLETE OUTPUT FLOW

#### NEVER USER OUTCOME

**EVER USER COMPLETE OUTPUT FLOW** **NEVER USER OUTCOME**
|  | No Interaction with the program | took a referral from Aapi | Took FP at doorstep | Total |
| --- | --- | --- | --- | --- |
| Never User | 43,623 | 7,498 | 5,469 | 56,590 |

#### NEVER USER COMPLETE OUTPUT FLOW

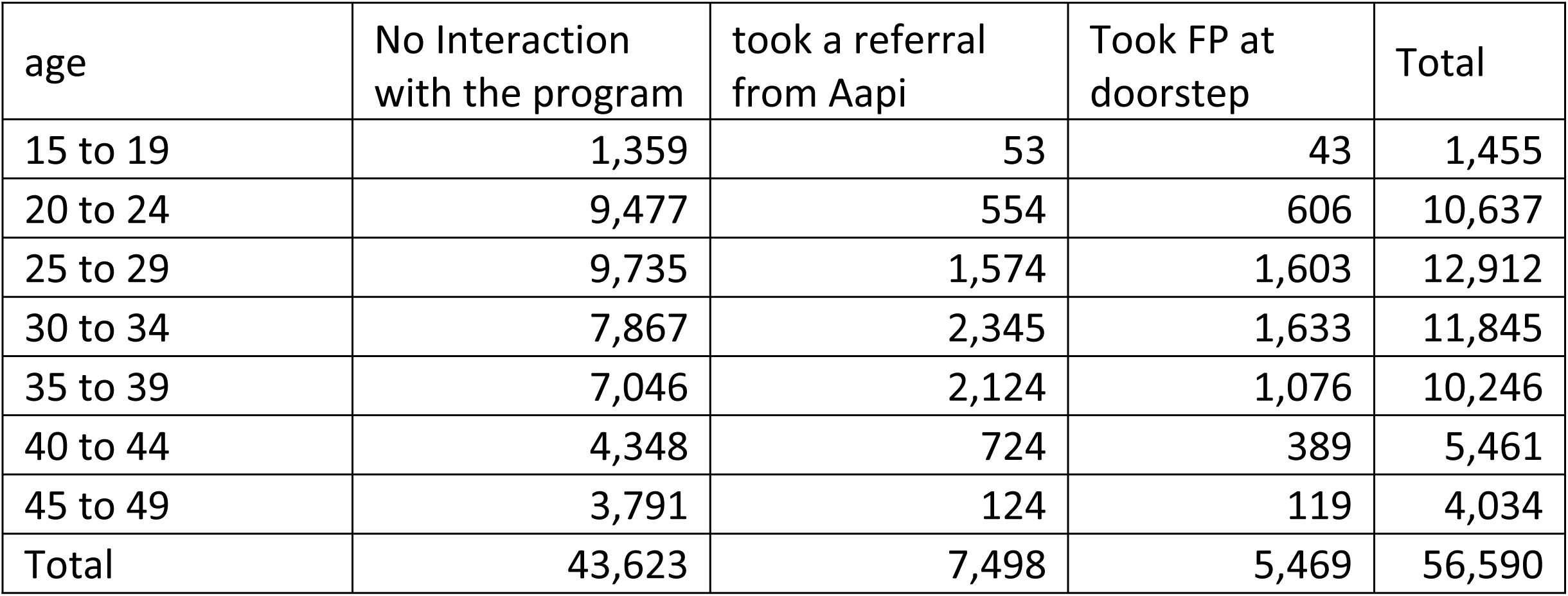

### NEVER USER COMPLETE OUTPUT FLOW

#### OVERALL IMPACT OF THE PROGRAM ON CONTRACEPTIVE USE

|  |  |
| --- | --- |
| Existing Users at Registration | 35,777 (36%) |
| Total users supported in using FP (continuing existing + new users) | 34,395 |
| New Users encouraged to start FP by Aapis | 12,967 |
| Existing Users served by Aapis | 19,012 |
| Existing users Converted from Short-Term to LARC Methods | 7,289 |
| Existing User opting to continue without Aapi support | 16,765 |
| Total Users after intervention (assuming continuation of users opting to use FP on their own) | 48,744 |
| CPR increase | 12.7% |

### NARRATIVE WRITE-UP

A total of 99,956 married women of reproductive age were registered during the study period. Overall, 34,395 women (34.4%) accessed family planning services through program-supported pathways, while 65,561 (65.6%) reported no program-linked service interaction. Among those who engaged with the program, service contact was distributed between doorstep provision by Aapis (15,772; 15.8%) and Aapi-facilitated referrals to health facilities (18,623; 18.6%). This overall pattern of engagement provides the context for understanding how service uptake varied by baseline contraceptive status, age, service pathway, and contraceptive method mix.

Women’s baseline contraceptive use status strongly structured their interaction with the program. Among women classified as current family planning users at registration, just over half (53.1%) accessed services through the program, split almost evenly between doorstep provision and referrals. Engagement among ever users was lower, with 31.9% accessing services, predominantly through referrals rather than doorstep delivery. Never users showed the lowest level of engagement, with only 22.9% accessing any program-supported service and more than three-quarters reporting no interaction. This gradient indicates that program reach was highest among women already connected to family planning at baseline, while initiation among women with no prior contraceptive experience constituted a smaller share of service delivery.

Across baseline user categories, program-linked service uptake was concentrated within the central reproductive age groups. Engagement peaked among women aged 30–34 years, where approximately 45% accessed services through either doorstep provision or referral, and remained high among women aged 25–29 and 35–39 years. Uptake was consistently lower among women aged 15–19 years and 45–49 years, regardless of baseline contraceptive status. Among never users, first-time uptake was largely concentrated in the 25–34 age range, whereas among current users, engagement extended through the late thirties, reflecting continued demand for spacing or limiting.

When examined by baseline contraceptive status, distinct method-use profiles emerged. Current users continued to rely substantially on short-term methods but also demonstrated considerable uptake of injectables and long-acting methods through referrals, including documented transitions from short-term methods to long-acting reversible contraceptives. Ever users exhibited a stronger skew toward injectable and long-acting methods relative to short-term methods, consistent with re-entry into family planning services after prior discontinuation. Never users overwhelmingly initiated family planning using short-term methods, with limited uptake of long-acting or permanent methods during the study period.

At baseline, 35,777 women (36.0%) were classified as current family planning users. Over the implementation period, the program documented family planning use among 34,395 women, including 19,012 existing users who continued use with program support and 12,967 women who initiated family planning as new users. In addition, 7,289 existing users were reported to have transitioned from short-term methods to long-acting reversible contraceptives. A further 16,765 women were reported to have continued family planning use without direct program interaction. When these groups were combined, the total number of family planning users following the intervention period was estimated at 48,744, corresponding to a 12.7 percentage-point increase in contraceptive prevalence relative to baseline, based on program-reported data.

At the time of registration, 7,289 currently using women (37%) had transitioned from short-acting reversible contraceptives, such as condoms, pills, or injections, to long-acting reversible contraceptives (LARCs) including IUCDs, implants, or tubal ligation. New user recruitment peaked around 12 months into the intervention suggesting saturation of potential users.

Across the analysis, consistency was observed between baseline contraceptive status, age profile, service delivery pathway, and method mix. Community-based doorstep provision primarily supported initiation and continuation of short-term methods, while referral pathways enabled access to clinically administered and long-acting methods. Engagement intensity and method complexity increased with prior contraceptive experience and age, suggesting internal coherence between program design and observed service utilization patterns.

### REGRESSION ANALYSIS FOR PREDICTORS OF FP SERVICE UPTAKE AMONG NONUSER AFTER EXPOSURE

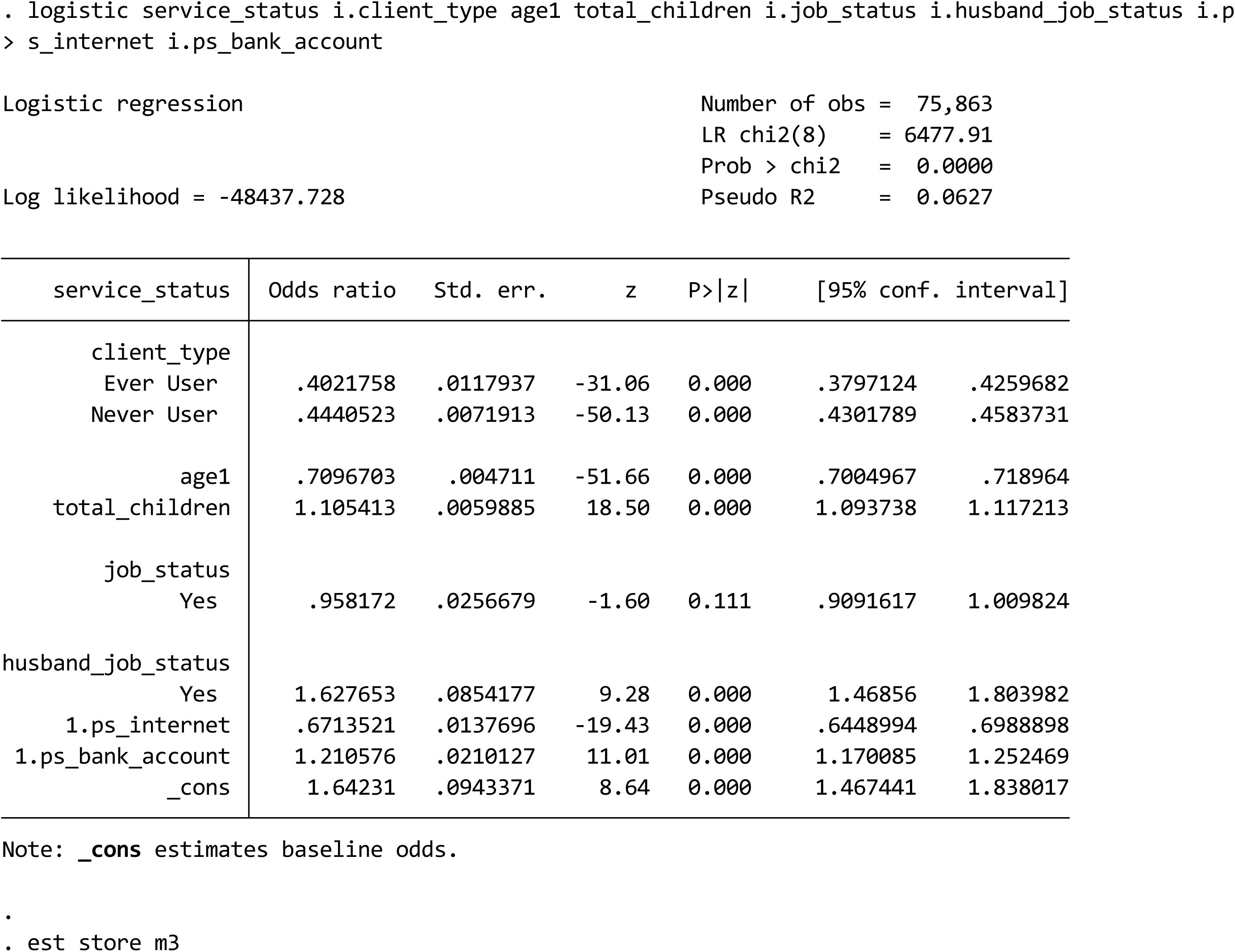

We estimated a multivariable logistic regression model to assess factors associated with service status (binary outcome). The model included women’s age, fertility composition (number of living sons and daughters), client type, urban wealth quintile, women’s employment status, husband’s employment status, and child immunization status. All categorical variables were entered as indicator variables with clearly defined reference categories.

| Variable | Category | AOR | 95% CI |
| --- | --- | --- | --- |
| **Age group** | 15–19 (ref.) | 1.00 | — |
|  | 20–24 | 1.16 | 0.94 – 1.44 |
|  | 25–29 | **1.34** | **1.09 – 1.66** |
|  | 30–34 | **1.39** | **1.12 – 1.71** |
|  | 35–39 | 1.19 | 0.96 – 1.48 |
|  | 40–44 | 0.86 | 0.68 – 1.09 |
|  | 45–49 | **0.57** | **0.40 – 0.81** |
| **Number of sons** | None (ref.) | 1.00 | — |
|  | One son | **1.45** | **1.35 – 1.55** |
|  | Two sons | **1.71** | **1.59 – 1.84** |
|  | Three sons | **1.82** | **1.66 – 2.00** |
|  | Four or more sons | **1.76** | **1.55 – 2.00** |
| **Number of daughters** | None (ref.) | 1.00 | — |
|  | One daughter | **1.31** | **1.24 – 1.39** |
|  | Two daughters | **1.47** | **1.37 – 1.57** |
|  | Three daughters | **1.56** | **1.43 – 1.70** |
|  | Four or more daughters | **1.26** | **1.12 – 1.41** |
| **Client type** | Reference group (ref.) | 1.00 | — |
|  | Ever user | **0.34** | **0.31 – 0.37** |
|  | Never user | **0.39** | **0.38 – 0.41** |
| **Urban wealth quintile** | Quintile 1 (poorest, ref.) | 1.00 | — |
|  | Quintile 2 | **0.71** | **0.66 – 0.76** |
|  | Quintile 3 | 0.98 | 0.86 – 1.13 |
|  | Quintile 4 | 1.12 | 0.92 – 1.37 |
|  | Quintile 5 (richest) | **1.75** | **1.46 – 2.10** |
| **Woman employed** | No (ref.) | 1.00 | — |
|  | Yes | 1.04 | 0.96 – 1.11 |
| **Husband employed** | No (ref.) | 1.00 | — |
|  | Yes | **1.41** | **1.19 – 1.66** |
| **Child immunized** | No (ref.) | 1.00 | — |
|  | Yes | **0.77** | **0.64 – 0.92** |
| **Constant** | — | **0.62** | **0.45 – 0.86** |

#### Insignificant Covariates

Several covariates were not independently associated with service uptake after adjustment. These included women aged 20–24, 35–39, and 40–44, women’s own employment status, and urban wealth quintiles 3 and 4. The lack of association suggests that marginal differences in age outside the core reproductive years and women’s labor force participation do not meaningfully influence service uptake once fertility composition and household characteristics are accounted for.

#### Fertility Composition, Family Completion, and Service Uptake

The number of living sons emerged as one of the strongest predictors of service uptake, exhibiting a clear and monotonic gradient. Relative to women with no sons, those with one son had 45% higher odds of service use, rising to nearly 80% higher odds among women with three or more sons (all p < 0.001). This stepwise increase strongly reflects a family completion dynamic, whereby engagement with services increases once households perceive that their desired family composition—particularly the attainment of sons—has been achieved. The magnitude and consistency of the association indicate that service uptake is not simply a function of parity but is closely tied to culturally defined reproductive goals. Women appear more willing or able to engage with services once son-bearing expectations are fulfilled, suggesting that service use is conditioned by perceived reproductive security rather than biological need alone.

In contrast, the number of living daughters was also positively associated with service uptake, but the pattern was weaker and less consistent than that observed for sons. Compared to women with no daughters, those with one to three daughters had higher odds of service use; however, the association attenuated among women with four or more daughters. Interpreted through a family completion lens, this pattern suggests that daughters contribute to service uptake primarily by increasing overall parity rather than signaling completion of fertility intentions. In settings where sons remain central to family formation ideals, higher numbers of daughters may indicate unfinished reproductive goals, resulting in weaker or less stable engagement with services. The divergence between the effects of sons and daughters thus underscores a gendered threshold of family completion, beyond which service uptake becomes substantially more likely.

---

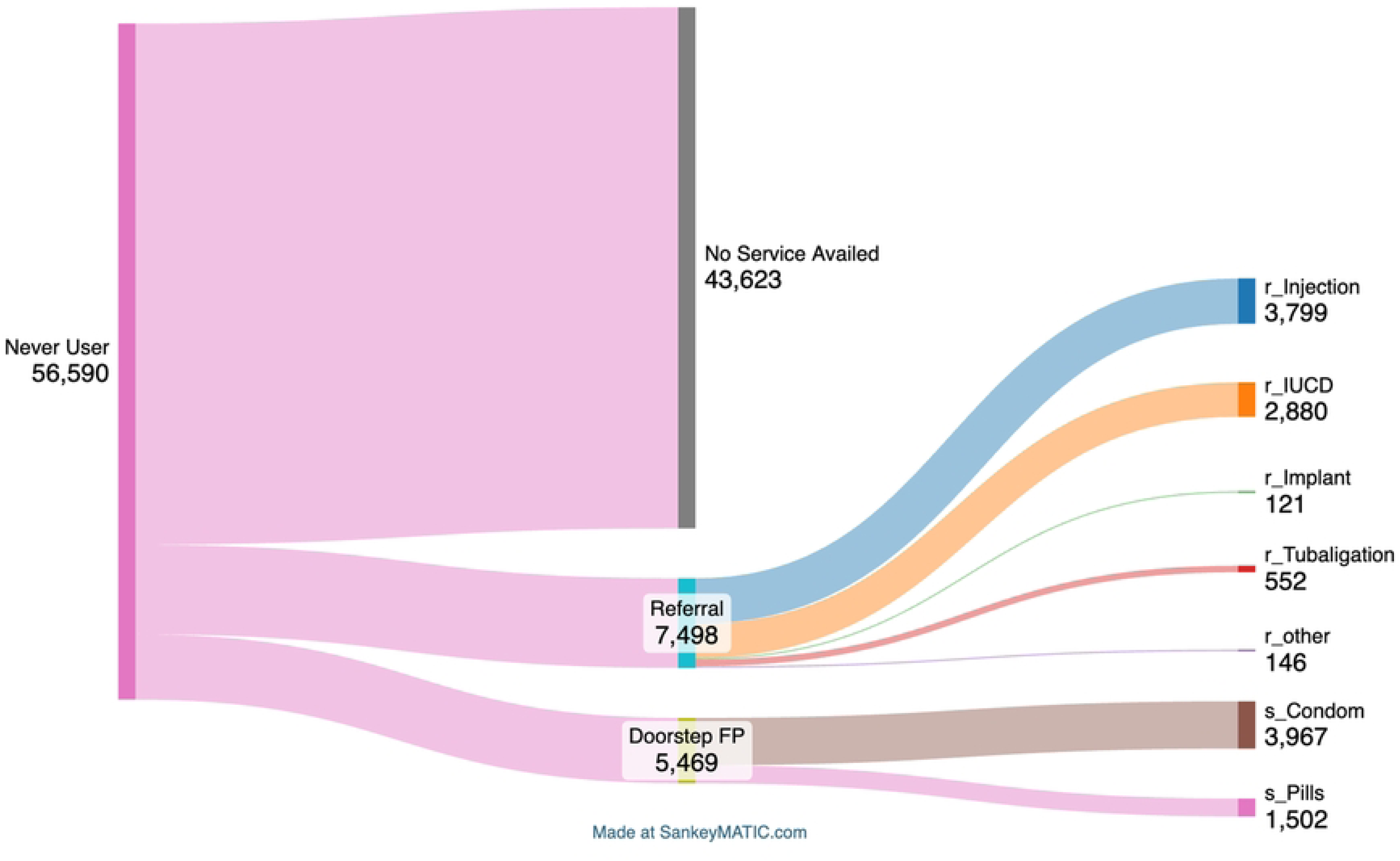

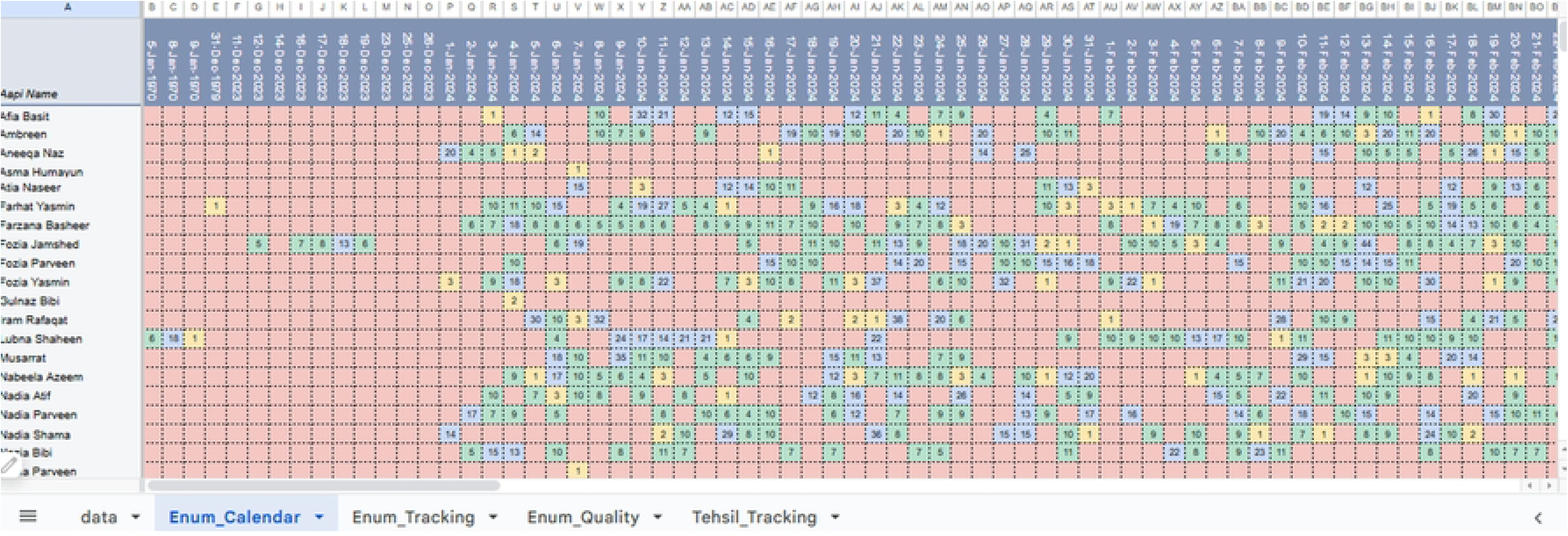

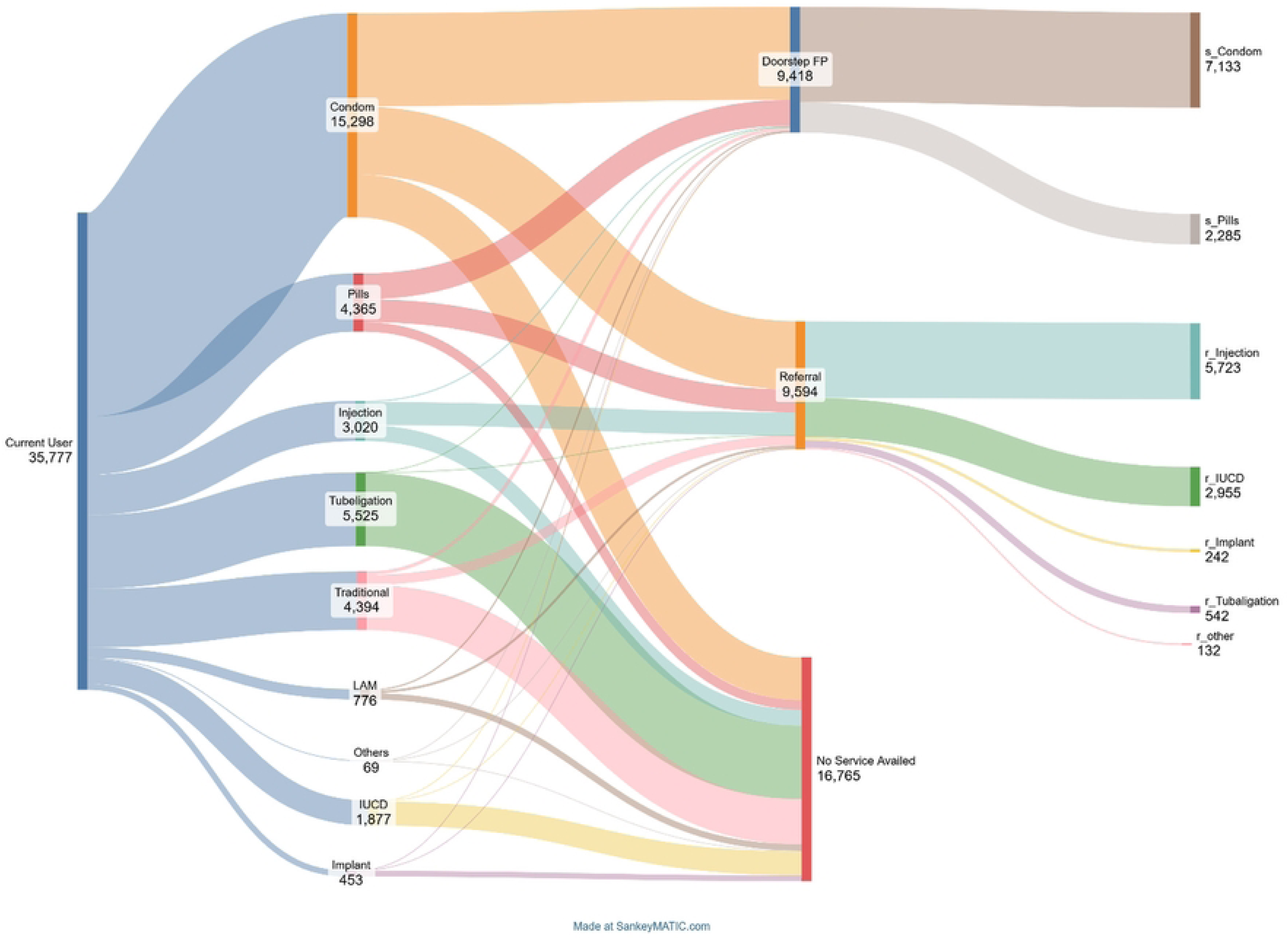

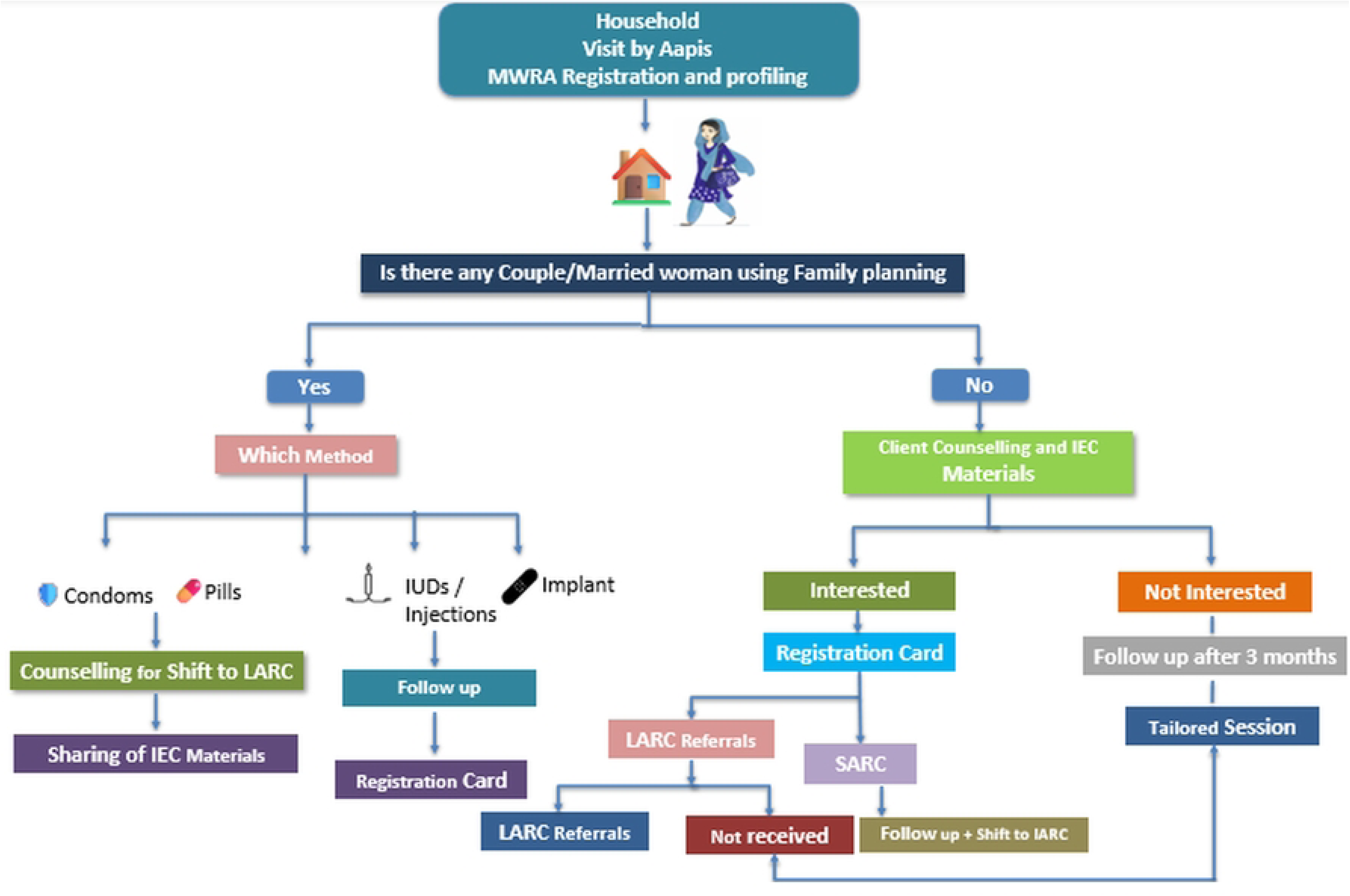

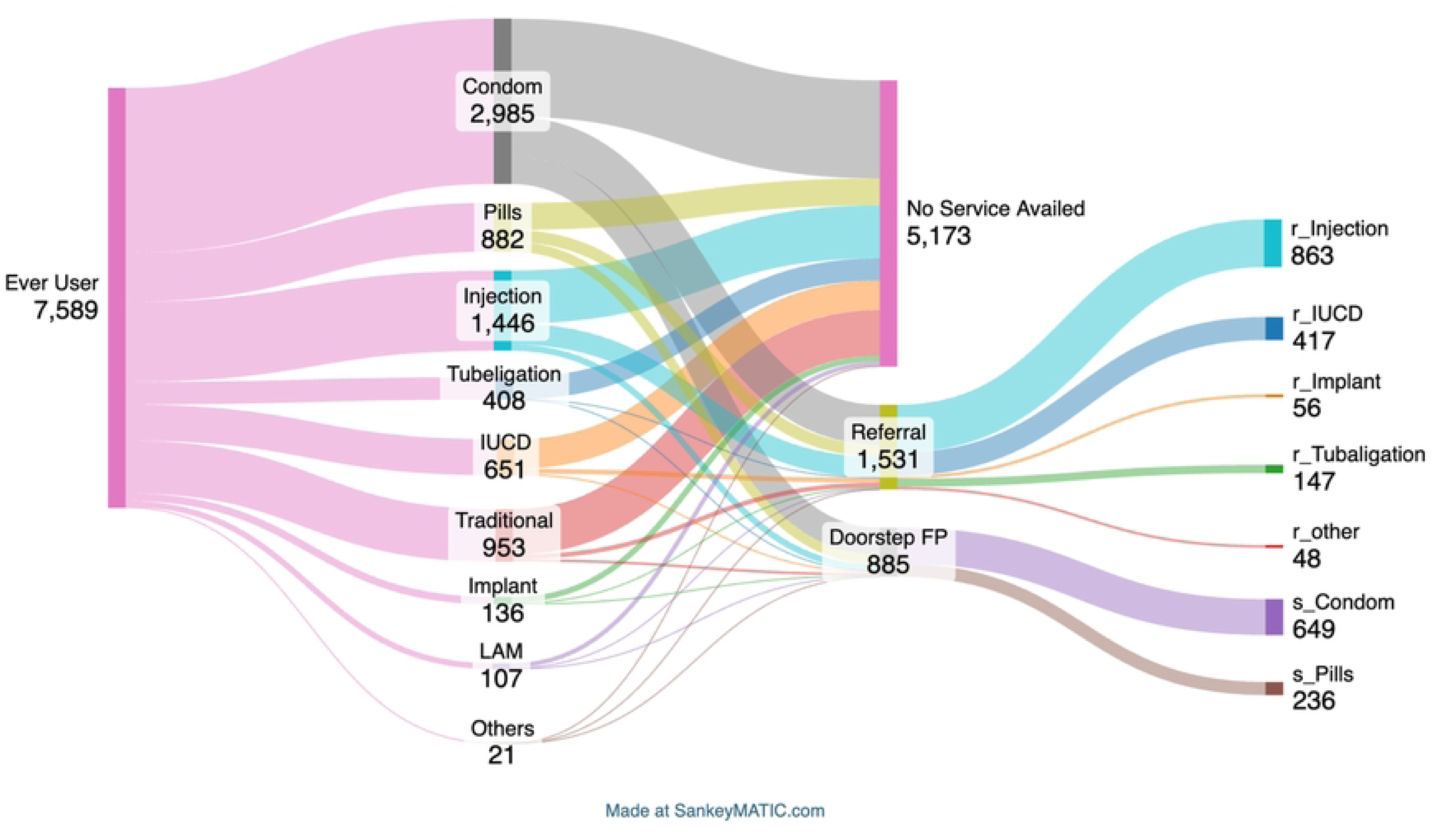

